# Telehealth use for sexual and reproductive health promotion and care during the early phase of COVID-19 pandemic: A descriptive-interpretive qualitative study of healthcare providers’ perspectives and experiences in Western - Central New York State

**DOI:** 10.1101/2024.05.02.24306759

**Authors:** Sadandaula Rose Muheriwa-Matemba, Danielle C. Alcena-Stiner, Alexander Glazier, Natalie M. LeBlanc

## Abstract

Telehealth emerged as an option for the provision of sexual and reproductive health (SRH) care and promotion during COVID-19 pandemic restrictions. However, studies are limited on the perspectives and experiences of healthcare providers (HCPs) practicing in the Western-Central region of New York State. This qualitative interpretive study explored the perspectives and experiences of HCPs with telehealth use for sexual and reproductive health promotion including counselling, testing, care and treatment for HIV infection and other sexually transmitted infections (STIs), in Western New York State. Ten HCPs participated in semi-structured in-depth interviews from October 2019-February 2021. These providers were predominately white, female, ranged in years of clinical experience (1-30 years). The narratives revealed three major themes: 1) healthcare providers’ perspectives of telehealth use, 2) healthcare providers’ experiences with telehealth use for SRH promotion and care, and 3) determinants of telehealth implementation. Though all providers reported an increase in the use of telehealth, experiences in the delivery of telehealth varied especially for sexual and reproductive health services. Some providers reported having more time to consult with patients because of a decrease in patient load which freed up time to engage with patients. Others reported technological limitations among some patients which impacted care. Strengthening telehealth-based sexual health promotion will serve to address efforts toward ending the HIV epidemic, reducing other STIs, and ensuring consistent access to contraception. To effectively implement telehealth findings, suggest a need to ensure adequate technological resources for patients, and a need to increase HCPs’ comfort to engage patients in sexual health conversations via telehealth.

## Introduction

Public health responses to the COVID-19 pandemic were unprecedented in modern times regarding its impact on healthcare delivery [1, 2]. Quarantines, institution-wide closures, and reallocation of resources towards emergency services compromised routine sexual and reproductive health (SRH) services [3–5]. Global disruptions in HIV prevention and care were associated with 650,000 HIV-related deaths [510 000–860 000] and 1.5 million [1.1–2.0 million] new HIV infections in 2021 [6]. Moreover, a 2022 scoping review by VanBenschoten et al [7] revealed 86% of SRH clinicians and stakeholders in 29 countries reported that patients had much less access to SRH services due to the pandemic. In the United States (US) the report identified an 80% decrease in HIV preexposure prophylaxis (PrEP) prescriptions. Other US studies reported a stark increase (33%) in maternal deaths after March 2020, albeit some resulted from direct viral infections, others have been attributed to the disrupted health care systems [8]. More than half of clinics canceled or postponed contraceptive visits, and 21% of SRH clinics in the South and Midwest parts of the country closed per COVID-19 pandemic mandates [7]. In addition, electronic survey results from a California-based sexually transmitted diseases (STDs) control branch found sharp declines in reportable bacterial cases of chlamydia (31%), late syphilis (19%), primary/secondary syphilis (15%), early nonprimary non-secondary syphilis (14%), and gonorrhea (13%) between 2019-2020 (9).

As the COVID-19 response intensified, telehealth services emerged as an option for accessing health care services, particularly those services considered non-essential, in order to maintain healthcare delivery in the US and globally [10, 11]. Telehealth, defined as the use of telecommunications and information technology including computers and mobile devices to provide access to health assessment, diagnosis, intervention, consultation, supervision, and information virtually [12–14], enabled patients to access support and clinical care from any location. E-health refers to health services and information delivered or enhanced through the internet and related technologies [14, 15]. Telehealth in combination with e-health helped HCPs to track and manage the health conditions of patients remotely to mitigate COVID transmission [16, 17]. An internet-based study among cisgender women in the US found that telehealth helped to fill in some new SRH service gaps [11]. Due to the pandemic, 24% of women using contraception switched to a telehealth appointment to have prescriptions refilled. Telehealth has been reported to be patient-centered, conducive to self-quarantine, and protecting patients, clinicians, and the community from exposure to infection [10]. More frequent check-ins through telehealth and remote patient monitoring also helped HCPs identify some complications faster, leading to fewer hospitalizations and emergency room visits [16, 17].

Telehealth service delivery during COVID-19 pandemic restrictions has been offered in different ways with live video teleconferencing being the most popular modality [18, 19]. Other modalities included store-and-forward technology which is the transmission of a patient’s medical information to a healthcare provider at a distant site [20], remote patient monitoring such as telehealth coverage of intensive care units, and mobile health applications including text, and email [16, 21]. In the US, most facilities have been reported to use a combination of telephone and video for telehealth visits [18, 19]. Many HCPs also used Health Insurance Portability and Accountability Act (HIPAA) compliant applications to conduct virtual telehealth visits, which assured patient privacy [19].

In the US, insurable services and processes, health policy, and practice may vary regionally. Therefore, HCP perspectives across state and county levels are needed to address the heterogenous subtleties in telehealth utilization for SRH promotion. Moreover, the experiences of HCPs in Western-Central New York State in their provision of SRH promotion and care during the COVID-19 pandemic has not been well documented. When New York State emerged as one of the epicenters for COVID-19 in 2020, SRH services were significantly reduced with nonessential services closing as of March 22, 2020, while hospitals around the state were encouraged to prioritize their resources to address the pandemic [4]. This impaired facility-based services and in-person health visits. Nevertheless, HCPs were encouraged to promote sexual health, conduct a complete sexual health history, and assess susceptibility to sexually transmitted infections (STIs). An advisory from the New York State Department of Health also offered strategies for healthcare providers and community-based organizations to support sexual and reproductive health and prevent STIs during COVID restrictions [22].

Understanding the experiences of HCPs’ delivery of SRH services in tandem with the uptake and use of telehealth during COVID-19 pandemic is warranted. Such insights can inform the suitability of telehealth for sexual and reproductive health promotion and care, and implementation needs for sustainability of this modality in Western-Central New York.

## Methods

This study used a qualitative descriptive-interpretive design [23] to understand the perspectives and experiences of HCPs with the use of telehealth for SRH promotion and care during COVID-19 pandemic. A qualitative descriptive interpretive approach allowed the investigators to address complex experiential sexual and reproductive health promotion questions during the COVID-19 pandemic, while understanding practical outcomes and underlying experiences. Researchers wanted to go beyond reporting the findings and interpret what the findings meant in the study context [24]. This approach allowed the advancement of knowledge and experiences surrounding SRH support during COVID-19 pandemic without sacrificing methodological integrity that long-established qualitative approaches provide [24]. Semi-structured in-depth interviews were conducted with 10 healthcare providers who had previously been interviewed about sexual health promotion and couple-centered approaches. This sample size was determined by saturation. These providers were a subset of this group and were practicing in women and adolescent health clinics, HIV/STI testing and care, and family medicine settings. Thus, the participants had previously interacted with the researcher in the first phase of the study. These healthcare providers were recruited from October 2019 to February 2021 using maximum variation purposive sampling [25]. The procedures and the analyses of this study were approved by the University of Rochester Institutional Review Board.

Active and passive recruitment strategies were used, which included circulating study flyers via emails; university research listservs and posting flyers in the health care facilities and on social media; and meeting providers at department or facility grand rounds to discuss the research. Subjects were asked to either contact the study team directly or complete a screener form in REDCap to assess their eligibility. Eligibility included being over 18 years old, speaking and understanding English, and being a licensed clinician practicing in New York State with one or more years of experience in sexual and reproductive health. All eligible participants were provided with an information letter that contained study details. Upon reading the information letter in REDCAP, providers indicated their desire to participate in the study, and they all provided written consent. Interested participants were asked to provide their socio demographic and contact information. Upon receipt of the notification of completion of the REDCap survey, the study staff followed up with a phone call and scheduled the interview. None of the participants who were sampled in this subset of the sample refused or dropped out during the interview. The interviews were conducted by the last author NML, a female PhD prepared Assistant Professor and an interdisciplinary nurse researcher, with over 20-year career in public health, nursing, and health research. NML uses multi-method qualitative/quantitative approaches to inquiry, and investigates the ecological, cultural, and systemic factors (social determinants of health) that influence health and wellness outcomes. As a public health specialist, NML is able to critically assess health issues from both public health and clinical perspectives globally and domestically. As a nurse researcher, NML seeks to address and investigate determinants of health disparity, assets within these factors that can be leveraged toward achieving health equity to inform intervention implementation. At the beginning of each the interviewer re-introduced herself and the purpose of the study and verified their interest to participate in the study. All conducted via Zoom, in a private room with only the participant present.

The individual in-depth interviews were guided by a semi-structured interview guide which was piloted on one participant and repeated on another one participant after making the corrections to the interview guide. Each semi-structured in-depth interview concentrated on the experiences of the participants with sexual and reproductive health promotion during the COVID-19 pandemic. This report is focused on the use of telehealth. Participants were asked the following main questions (with follow-up prompts which are note listed):

1. Describe the impact of COVID-19 pandemic and quarantine has had on your personal practice in sexual health promotion with patients.
2. What changes have you made to your personal practice in sexual and reproductive health, HIV/STI prevention, and/or care promotion during the COVID-19 pandemic?
3. Describe your experiences with using telehealth during this time including what worked well for you as a provider and for your patients?

The interviews lasted 30-45 minutes and each participant was paid 30 US Dollars for their time. All interviews were recorded with HIPAA compliant University of Rochester Medical Center Zoom and iPad. Field notes were also taken to capture the necessary contextual information. Data were transcribed verbatim by an outsourced transcription company.

### Data Analysis

A thematic analysis involved an inductive iterative approach in the generation of codes and themes and subthemes [26]. All themes and subthemes were generated from the data. Upon transcription, the first, second and senior author read each transcript at least 3 times line-by-line to achieve immersion into the data and obtain a sense of a whole while identifying the underlying concepts and clusters of concepts [26, 27]. These authors read each transcript, highlighting text that was salient and appeared to describe perspectives and experiences with SRH promotion and care; and using telehealth during the COVID-19 pandemic. Memos noted initial impressions, as well as keywords or phrases that captured the perspectives and experiences of health providers.

Memoing allowed a reflection on findings to deepen our analysis, generate new ideas and interpret and communicate our findings. Memoing also assisted in making conceptual connections from raw data to those abstractions that explained the use of telehealth for sexual health promotion during COVID-19 pandemic (28). Via an iterative process to create a coding schema that reflected critical thoughts and codes that came directly from the interpretation of the interviews. This process was facilitated by MAXQDA 2020 software [26, 27, 29].

After coding all transcripts, during peer debriefings the investigators examined each code and developed and refined descriptive statements for each code. This process led to combining some codes to form main themes and splitting others into sub-themes based on how different codes were related and linked to each other. These emergent themes helped to organize and group codes into meaningful clusters [30, 31], while keeping them broad enough to sort a large number of code [32]. Depending on the relationships between sub-themes the researcher combined and organized the more substantial number of subthemes into a smaller number of themes. Next, the definitions for each theme and sub-themes including the text segments that corresponded to each theme were documented in the coding book [33]. Themes and subthemes were then organized [29] to illuminate health care providers’ perspectives and experiences with SRH health promotion and telehealth use during COVID-19 pandemic. Peer debriefing was conducted to ensure that there was agreement or reconciliation with the findings, coding, and thematic scheme. Later findings were presented to the participants for them to verify the results.

## Results

Table 1 shows the sociodemographic characteristics of the participants. The participants were predominately white, female, and ranged in years of experience (1-30 years). Narratives from participants revealed three major themes as relates to telehealth use for SRH promotion and care: 1) healthcare providers’ perspectives of telehealth use, 2) healthcare providers’ experiences with telehealth use for SRH promotion and care, and 3) determinants of telehealth implementation.

**Table 1.** Socio Demographic Characteristics of the Participants, (*N*=10)

| Sociodemographic Characteristic | *M* | *Range* | *n* | % |
| --- | --- | --- | --- | --- |
| Age | 51 | 28-67 |  |  |
| Sex |  |  |  |  |
| Male |  |  | 4 | 40 |
| Female |  |  | 6 | 60 |
| Ethno-Race |  |  |  |  |
| Non-Latino Black/ African American |  |  | 1 |  |
| Non-Latino White |  |  |  |  |
| Latino |  |  | 1 |  |
| Other |  |  | 1 |  |
| Religion |  |  |  |  |
| None |  |  | 3 | 30 |
| Christian |  |  | 7 | 70 |
| Type of Health Profession |  |  |  |  |
| Medical Doctor |  |  | 5 | 50 |
| Nurse Practitioner |  |  | 2 | 20 |
| Nurse Midwife |  |  | 1 | 10 |
| Physician Assistant |  |  | 2 | 20 |
| No. of years in the Health Profession | 21.3 | 2 - 43 |  |  |
| No. of years the HCP has been licensed in the profession | 23.1 | 2 - 43 |  |  |
| Type of facility the HCPs worked |  |  |  |  |
| Public Clinic |  |  | 2 | 20 |
| FQHC |  |  | 3 | 30 |
| Private & Academic health facility |  |  | 3 | 30 |
| Hospital based clinic |  |  | 1 | 10 |
| Outpatient clinic and juvenile detention center |  |  | 1 | 10 |

Note: FQHC = Federally Qualified Health Center; HCPs= Healthcare providers

### Healthcare Providers’ Perceptions of Telehealth

Narratives revealed HCPs perception of telehealth especially use for SRH promotion and care. Participants reported their perceived benefits of using telehealth despite some limitations during inception. Their perspectives were in part, influenced by telehealth as a modality, their experience with telehealth uptake in response to the COVID-19 restrictions, and due to organizational considerations for the adoption of telehealth service. Perceptions were also influenced by patient capabilities to engage with the technology. One participant said: “…*the more you make it available and decrease the sort of limitations to access, uh, it seems to be more successful and widely used* (Nurse-Midwife, > 25 yrs.). Another participant said: …*it may improve access for people as long as you can work out some of the other logistics*….(PA, <10 yrs.).

Providers reported that telehealth use increased accessibility for HIV and other STIs during the COVID-19 pandemic. It also enhanced the provision of couple and family-based care.

> … it has become a little bit easier to talk to people together with telemedicine… somebody can kind of just casually join a visit, even if they’re not on the schedule, which is not something that’s possible, you know, in an office setting… that’s actually been kind of nice (PA, <5 yrs.).

Another participant added:

> At least in New York state, telehealth has been shown to not only increase testing to populations that weren’t previously reached by in-person …. but also, it seems to increase testing in sexual networks because you can have a kit and maybe you can have a second one and give it to your partner…I feel like all those things have sort of taken flight…(Midwife, >30 yrs.)

> Participants also perceived that telehealth use for patients can ease engagement with the healthcare system, and potentially address stigma arising from HIV/STI prevention and care.

One participant said:

> …. because you [patients] can request a kit and not have to come in contact with a health care personnel. This overcomes part of the stigma associated with seeking care for STIs because you’re able to do something online. It can be anonymous, or even if it may not be, but it sort of takes away that step of having to go somewhere in person (MD, >25 yrs.).

### Experiences with telehealth use

For most providers, the emergent use of telehealth and e-health was in the context of health facility’s responses to the COVID-19 pandemic. Providers’ perspectives and experiences with telehealth, especially in the context of sexual health (including HIV/STI testing and care) varied and was in part influenced by how health facilities responded to organizational accommodations to COVID restrictions. Statewide COVID-19 restrictions required healthcare facilities to re-assess their provision of what was considered non-essential health services. This included routine sexual and reproduction care including HIV and STI prevention, diagnosis, and treatment services. Re-assessment entailed either healthcare services being exclusively redirected to address the health needs resulting from COVID-19 pandemic; provision of essential health services in addition to COVID specific care; or exclusive modification of sexual health services to remote telehealth services with occasional triaged in-person follow-up visits. This modification resulted in some instances, halting, and slowing the provision of sexual health services. In other instances, services remained intact with the addition of telehealth.

> We pivoted to telemedicine visits, mail-in pharmacy, doing telehealth visits and STI testing by sending testing materials, swabs, instructions, and lab materials and requisitions to people at home so that they could collect specimens at home, drop them off in the lab. In other words, trying to make this as seamless as possible and continue what we consider essential services…. We also opened a COVID testing area that is still in operation (MD, primary care, > 40 yrs.).

> Healthcare providers also reported eventual adoption of telehealth due to experiencing a slow down or ceasing of SRH services to focus on COVID-19 prevention and care activities. Some providers reported adopting telehealth services following a better understanding of COVID-19 transmission from an epidemiological and clinical perspectives.

> … we do pregnancy and then gynecological care. We were limited as to how many people we could bring in. Patients that were having gynecological problems, STDs or needed contraception, they kind of took a back seat to what was available as far as appointments. So, we were able to offer them tele-health that helped them access care (ARNP, women’s health, > 25 yrs.).

### Approaches to telehealth modalities

Providers reported on approaches to telehealth in the context of the agency’s capacity in tandem with the health facility’s focus on HIV/STIs prior to the onset of the COVID restrictions. Providers reported use of different approaches to telehealth use which included telehealth by appointment, walk-in video visits and other web-based modalities such as PrEP2Me.

> It’s really like making it readily available where you can meet people where they are. …in some places you can only do it by appointment, so that’s very restrictive…here, you can just call in and get, go through a visit at any time during the time the clinic is open. There are other places where it starts as kind of a web-based thing. So, you just submit your information and then they reach out to you directly. … (Nurse-Midwife, > 25 yrs.).

Provider reports indicated that video telehealth was the most preferred modality for telehealth visit as it allowed them to see and physically assess their patients.

> … I saw somebody recently…we asked how, how the person was doing, and they said fine, but on the video, they’re crying. So, you know, on the phone, I wouldn’t see that…because you couldn’t hear it in the voice necessarily (PA, < 5yrs.).

Overall, the HCPs reported positive experiences with telehealth use. The participants also reported of how they found telehealth to be a feasible and acceptable modality of providing care during the pandemic. Positive experiences were related to provider ease and comfort not only with modality, but as well patient capability also to have sexual health needs met. There were cases where providers reported that telehealth facilitated the adoption of certain services like at-home HIV/STI tests, and patient capability to self-collect specimens. This modality helped with continuity, yet alternatives in the provision of sexual health services.

> I think the transition has gone reasonably well and, and people like the option of having some telehealth visits… telePrEP is probably a pretty good thing. And I think it may improve access for people as long as you can work out some of the other logistics (PA, <10 yrs.).

Another participant also said:

> The people I’ve talked to have, seem to like it [telehealth]…that includes HIV testing and, genital and extra genital swabs. And people are doing their own rectal swabs and throat swabs, and they’ve learned to do it and are comfortable with it. I was, um, pleasantly surprised at the comfort level and they accept it (MD, >25 yrs.).

### Challenges of Using Telehealth

Providers reported challenges they faced with using telehealth. These included: loss to follow up, lack of opportunity to examine the patient, internet interruption, scam calls that made patients not respond to real phone calls for the hospital visit, lack of privacy, lack of time management and seriousness by the clients.

### Reduction in patient panel

While some providers reported an increase in patients who normally would not engage, other providers reported that in-person restrictions resulted in a reduction in patient volume and patients’ loss to follow-up.

> …We did lose about 10% of our 1000 patients though who drifted away and have been a little harder to reconnect. But overall, I think it certainly was preferable to not doing anything. And we made an effort to stay connected with, uh, with people (MD, > 25 yrs.).

As some providers report and increase in HIV tested via telehealth other providers reported a reduction in patients as a result in restrictions on their practice. One participant said: …*there was probably decreased access, not because we weren’t open, but because patients weren’t coming in to get tested* (MD, 10 yrs.).

### Limited physical assessment

Providers reported that telehealth use, though engaging of patients, there wasn’t an opportunity to conduct a physical assessment. Providers deemed this to be one of the essential aspects of a sexual health promotion visit. Not having an opportunity to physically examine the patient when the patient posed as a limitation of telehealth use.

One participant said:

> Yeah, sometimes telemedicine tends to be, um, more challenging because you can’t examine a person and some people do need an exam, so that, you know, you do have to defer that to a different setting, but, um, it really is well suited to sort of preventive care or well care (Nurse Midwife, >25 yrs.).

> … there are just some situations where you can’t do it by video. When you need to touch the patient and you can’t. Or listen to the patient’s lungs… if they had a rash… We would attempt to set them up for a video appointment because you can describe a rash as best you can. But I mean, that visual is what makes the diagnosis. (MD, 10 yrs.)

### Technology landscape

Internet interruption was another challenge that the participants experienced particularly with those clients of low socioeconomic status.

> … sometimes it just doesn’t work… they just don’t have the Wi-Fi, and it’s not very good. And so we have to end up turning off the video to hear what the person’s saying…lack of internet or lack of any type of video resource pose the challenge (PA < 10 yrs.).

The participants also reported on challenges posed in part due to lack of familiarity with the technology for both providers and patients.

> …it was like a big adjustment to me, …more so a challenge, I guess, like for the patients, just understanding that you know, this is a telephone appointment, or this is a video appointment and getting them, the proper resources that they needed to, be prepared for a telephone or video appointment (MD 10 yrs.).

Providers also reported that patients at times would not answer hospital calls thinking that they were scams. In response, some health facilities introduced phone applications that helped to overcome this challenge.

> …we use, Doximity… it’s targeted towards clinicians… the phone number shows up, which is really important on their phone. So now they’re answering our phones, and then basically click through a series of allows… it’s HIPAA compliant. (PA, <10 yrs.)

### Lack of privacy

Providers reported concern with privacy during telehealth consultations. Providers reported that patients were not always in a private location during the time of the telehealth visit consults.

> So, you do get into these dicey issues, and even though when people say that they’re in a safe space, I sometimes wonder and, uh, you know, confidential space, uh, you know, when you’re out in public or what have you, you know, how confidential is it? (PA, <5 yrs.).

Although some providers perceived couple or family-based telehealth visits favorable, others had some concerns. Some providers expressed uncertainty of whether they addressed patients’ needs if they sensed someone else like a family member was present in the home during the telehealth visit.

> …here we separate partners for most of the visit, to ensure people are comfortable and sharing confidential information, because here we have a lot of privacy rules. But when you’re doing telehealth you can’t, um, you’re not always sure that you’re getting the full story, um, because there’s no sort of confidentiality (Nurse Midwife, > 25 yrs.).

### Determinants of telehealth implementation

Implementation of telehealth was determined by providers experiences of the organizational capacity to facilitate telehealth, the healthcare provider’s comfort to manage sexual health dynamics, and technology landscape.

### Organizational capacity to implement telehealth

Organizational capacity spoke to providers reports of sustainability of telehealth as part of healthcare. Providers reported on prior knowledge and experiences with telehealth Providers particularly those with experience of using telehealth prior to the pandemic reported positive experiences of using telehealth and the ease of rolling out telehealth for sexual health promotion. One participant said: *“Well, the telemedicine works well. We have been experimenting with that a little bit, prior to that [COVID-19 pandemic]. And so we had a little bit of a foundation and then just turned it, …”* (MD > 25 yrs.).

Organizational capacity to implement telehealth rested on several factors as reported by providers. One factor was the ability to reimburse for the provision of healthcare services via a telehealth visit. Providers reported that the insurance companies would only reimburse for the video-based telehealth visits and not phone-based visits. Providers reported that the insurance companies’ reluctance to reimburse telehealth visits by phone was a major barrier that made care for some patients inaccessible. Some patients do not have smartphones or WIFI to be able to have a video call telehealth visit.

One participant said:

> …I don’t think it’s fair to assume that every patient has an iPhone or a smartphone

> …there are patients that need only phone visits, and cannot afford video calls. And to make primary care sustainable, you really have to be able to care for patients in multiple ways. So, to assume that all patients have access to technology is inequitable (ARNP >10 yrs.).

Another participant added:

> … obviously, we want video, it’s better, you get better quality, but if the person can’t do it, what are you going to do? And why should the health center take the hit financially? Because we’re meeting the patient with the technology they have. It’s not gonna be sustainable to do phone visits based on what many of the payers are reimbursing (PA <10 yrs.).

Providers reported a number of personal and organizational strategies that facilitated implementation of telehealth for SRH promotion and care. HCPs reported of their adaptation to pandemic restrictions and understood that organizational financial loss may be inevitable. Other providers reported that instead of physicians or nurse practitioners providing all services, the organization leveraged an interdisciplinary clinical team and optimized sexual health service provision by nurses and social workers.

> So, ultimately primary care will not survive if it’s not paid for adequately by insurers. … we have always worked to use things like nurse care management as part of PrEP and part of STI care. … moving that forward a little bit and really using, like, nurse phone calls with patients instead (ARNP >10 yrs.).

Another participant added:

> …having our social worker who follows up patients has facilitated the telehealth. She typically does follow-ups to make sure that the patient’s partner has been treated. …Um, but she also follows up in the meantime too, especially if the patients tend to miss their appointment (PA, <10 yrs.).

## Discussion

Our study on exploration of HCPs’ experiences with SRH promotion during the COVID-19 pandemic revealed telehealth’s major impact on care delivery across the Western and Central regions of New York state. During the pandemic, sexual health care was not regarded as essential [3, 34, 35]. HCPs transitioned from in-person visits to new appointment modalities and use of mail-order pharmacies, and home STI /HIV testing. This significant shift in care provision for clients exposed facilitators and barriers to telehealth use among HCP in SRH. General barriers were a lack of provider comfort to manage sexual health dynamic and socioeconomic health disparities. These impediments have also been reported in previous studies [18, 19, 36] and are indicative of telehealth being a relatively new mode of care delivery. Telehealth was also perceived to reduce sexual health stigma. Findings indicated that with telehealth, people who would not seek health care due to anticipated stigma, associated with seeking care. Also, home-based testing may have helped increase STI screening within sexual networks and help linked people to care including PrEP care [37, 38]. This finding might explain why as testing for HIV and other STIs reduced significantly globally due to the COVID-19 pandemic (39), in Western New York State, the testing services increased and new STI diagnoses increased by 77% [22].

HCPs in this study reported telehealth benefits for easing access to patients, enhancing the provision of family and couple-based care, and increasing patient comfort, which can be foci for training current and future providers. Such training must be population specific and culturally responsive to client needs and perspectives while simultaneously provide modalities of telehealth care that are appropriate for HCP use in SRH. Despite telehealth being commended as a method for sexual health promotion, this study revealed some important determinants for telehealth implementation. Providers reported challenges to implementation that included: loss to follow up, limited physical assessment, internet interruption, and lack of privacy. Some loss to follow up could be addressed by client access to reliable internet service that is free or affordable, a device with video capabilities and audio/headphone features such as noise cancelation, or access to a safe space with privacy. Optimizing telehealth implementation also requires adjustment to new behavioral expectations for engagement such as looking at a camera for direct eye contact instead of the screen displaying the image of the other person [40]. A client may appear to be uninterested or not focused on the discussion when looking away, impacting the HCP’s interpretation of the client’s mood and affect. Yet, the observation may result from technical concerns related to camera and screen positions. HCP narratives also described limitation for assessing clients’ skin conditions for diagnosis, suggesting need for resources and guidance on clothing choice and lighting, as an environment influences appearance during virtual physical assessments. Perhaps clients need to be provided with telehealth resources that include the necessary equipment to maximize the telehealth visit [40].

Promoting telehealth etiquette can impact unintended interruptions and distractions that can include a choice of background images, filters, attire, or nontraditional visit in public locations. A client may willingly disclose elements of their environment or share that they are speaking candidly while others are present (on camera, or off-camera). However, HCP’s might be uncomfortable and distracted during the health care encounter; echoing the need for HCP to consider practice guidance that mutually reflects client’s input related to intended and preferred telehealth etiquette expectations. HCP’s perspectives in this study support other reports that suggest a need for “screen side etiquette” during telehealth assessment and care to optimize settings (e.g., visual, audio, etc.) and reduce interfering factors from the health care setting or client’s environment [41].

Collective HCP perceptions in the current study underscore the lack of infrastructure preparedness and training for broad use of the telehealth modalities described contribute to the field of research. Prior to the pandemic perspectives on the underutilization of telehealth training in primary care and specialty care for nurse practitioners (NP) and students in NP programs had been reported with the need to increase provider comfort, knowledge, and skills necessary for embracing telehealth at the forefront of health care [42].

The implementation of telehealth for sexual health promotion was facilitated by several factors. Organizational-level factors included the health facility’s capacity in tandem with the already existing plans to introduce telehealth prior to the onset of the COVID-19 pandemic restrictions. Narratives indicated that health facilities that had an already existing plan to implement e-health sexual health services, simply expedited their plan and strategically put in measures to help their clientele adapt to the emerging approach to remote sexual health care delivery. For some, the sudden surge in national use without established governmental and organizational guidance was more of a pandemic attribution than initiation of telehealth in practice. This suggests telehealth was underused and understudied until the COVID-19 pandemic, during which reduced regulations and increased payment parity facilitated a rapid increase in telemedicine consultation [50]. This underutilization and the limited studies regarding telehealth in specialty care may explain why some healthcare providers in this study reported their hesitancy and perceived lack of knowledge, demonstrating mistrust of its effectiveness and adherence to ethical standards and advocating for a combination of virtual and in-person care.

Findings also suggests the need for patient orientation to seeking sexual health considering that telehealth is currently being integrated into the health system. In contrast, agencies that had no existing facilities reportedly were challenged by initiating telehealth use for SRH services. Such that meeting typical and standard SRH needs slowed down or ceased altogether to focus on COVID-19 prevention and care activities.

Providers eventually within their roles began strategizing on how to meet patient’s health needs via telehealth and started slowly to implement telehealth services. Thus, the health providers’ attitude and knowledge of telehealth, and the cordial working relationship between physicians, advanced nurses, nurses, and social workers facilitated and enhanced the patients’ access to sexual and reproductive health care, vital elements in continuity of care [43].

The healthcare providers in this study, like in other previous studies [19] reported the challenges they faced with health insurance not willing to cover the cost when some modalities such as phone calls were used. It was under such circumstances that the healthcare providers’ understanding and willingness to meet the healthcare needs of the patients was essential. This gesture was commendable and aligned with the multifaceted patient-centered sexual healthcare delivery framework which Ohta, Ikeda and Sawa [44], Cuomo, Zucker and Dreslin [45], Gunaratne and Hansen [46] advocated for during the pandemic. Implementation of telehealth was made possible due to provider willingness to render sexual health services even when the facility was faced with profit loss due to reimbursement prohibitions the HCP narratives presented in this study and in other study reports [47].

Additionally, patients’ and HCPs’ understanding of telehealth and its necessity allowed telehealth to be doable, speaking to sustainability of this modality. Notably, 48 states including Washington, D.C., covered some degree of telemedicine in 2016 compared to only 24 states in 2005, based on American Telemedicine Association Coverage and Reimbursement report [48]. Therefore, telehealth reimbursement, although an increased priority for wide-spread enactment, was not a novelty due to the pandemic, as policies and logistics were previously established nationally. Despite increased national dialog, HCPs and patients were not well oriented to telehealth, and most may have needed more time to adjust, due to the rapid uptake of the modality warranted by COVID-19 and in-person restrictions [47]. Therefore, more research, provider and patient training, and orientation are required to develop better strategies and trust care in provision via telehealth service for sexual health promotion [49].

An interdisciplinary lens and subsequent cordial working relationship among the nurses, physicians and the social workers was also reported to facilitate the telehealth services. The nurses were reported to help with other visits that did not necessarily require the physician’s attention such as PrEP management and STI care, while the social workers helped by following up with patients who missed appointments and needed treatment refills, in addition to making sure partners of those diagnosed with STIs were treated. This finding is consistent with previous studies that reported telehealth physicians to be more dependent on their social systems, particularly nurses, than physicians working in-person [50]. During the pandemic, nurses have been reported to play a significant role in ensuring the visit is well coordinated and in ensuring a smooth workflow [50]. Thus, nurses are essential in the delivery of telehealth-based care.

Additionally, the narratives in this study showed that social workers are also critical to the implementation of telehealth for SRH promotion and care. Previous research suggest that social workers help to harness a range of skills and competencies of importance to the emerging needs of the patients during the pandemic including risk assessment, crisis management, advanced care planning, case management, systems navigation, problem solving, resource allocation, and community mobilization [51, 52]. These results show that a meaningful collaborative and mutually respectful interdisciplinary relationship is critical for achieving quality outcomes for patients and for the longevity of the telemedicine operation.

As in-person health services resume, health agencies must consider strategizing to maintain telehealth services in their facilities. Moreover, providers in this study, reportedly saw the merit for telehealth beyond COVID restrictions. Nevertheless, studies report differing views regarding whether telehealth is sustainable for delivering sexual and reproductive health services and that there are some considerations for planning integration of telehealth services. In a previous study by Yelverton et al [19], the participants questioned whether it was feasible for HIV testing services and PrEP to be provided entirely through telehealth and suggested a hybrid combination of remote and in-person delivery. The study also showed that participants questioned whether HIV home testing was done in a proper manner and stressed the need for in-person coaching of clients to ensure high-quality HIV testing and reliable results.

Hollander and Carr [10] report that telehealth needs to be prioritized when healthcare providers are planning health visits. They suggest that health facilities that have telehealth services need to leverage them. Systems lacking such programs can outsource similar services to physicians and support staff provided by multinational telemedicine and virtual healthcare companies. Hollander and Carr [10] and Yelverton et al [19] observe that the current major barrier to large-scale telehealth, is HIV/STI screening that can be overcome by proper coordination of testing and expanding testing sites. It is pivotal to educate future health care providers about these social determinants so that they can work to mitigate the resulting disparities and thereby improve the health of patients and their communities.

### Implications

This study has implications for research and practice. Public health and clinical efforts must acknowledge SRH promotion as an essential service during the pandemic. Findings suggests the need for all health care agencies offering SRH support to institute measures ensuring that sexual and reproductive health care and promotion is sustainable post-pandemic. This includes making sure that telehealth, mail pharmacy and the mailing of HIV and STI testing kits, which have been proven to be essential in enhancing the provision of sexual health services in Western-Central New York be adopted. This study can also help all HCPs to adopt the multifaceted patient-centered sexual healthcare delivery which was promoted in New York State, that helped patients to receive individualized and wholistic care. The results also show that a lot of studies are needed to further explore and find strategies to incorporate couple-based based approach to care and telehealth to make sure that patients receive comprehensive care and be able to reduce, HIV infections and other STIs.

### Limitations

Perspectives from HCPs who practice as medical doctors and physician-assistants used the term “telemedicine” when reflecting on changes in health care modalities, while others predominantly used the “telehealth” terminology. Although our findings focused on telehealth, expanding terminology to include e-health and m-health or even telemedicine may prove necessary for broadening determinants of utility and areas of training for implementation across the US and globally [14, 42]. Furthermore, the telehealth use for SRH promotion during the COVID-19 pandemic in this study has been reported from the perspective of healthcare providers only. Future research needs also explore the telehealth use for SRH promotion and care during COVID-19 pandemic from the perspectives of patients and service administrators.

## Conclusion

This study revealed perspectives emphasizing telehealth as a vital health care technology for continuity of care among clients facing impaired SRH service access, particularly in times when such services are not considered essential. As previous research shows that a slight reduction in HIV and STI screening may increase the rate of HIV infection by almost 8% [53], the increased use in telehealth for SRH promotion and care must be prioritized in research and intervention development. Such efforts can help to continuously meet the sexual and reproductive health needs of all people, achieve the goal to end the HIV epidemic, and reduce the rates of STIs in Western-Central NY and US at large.

## Data Availability

Data cannot be made publicly available due to patient confidentiality reasons, in adherence with the data access rules of University of Rochester IRB. Data can be accessed upon request from the following contacts: a. Office of the Vice President for Research, University of Rochester Research Subjects Review Board, 265 Crittenden Blvd., CU 420628, Rochester, NY 14642, Telephone numbers (585) 273-4127, (585) 276-0005 or (877) 449-4441. b. The PI Dr. Natalie LeBlanc at c. The corresponding author Sadandaula Rose Muheriwa-Matemba at

http://doi.org/10.791/DVN/EOEIFU

## Acknowledgements

Authors would like to thank all HCPs who participated in the study.

## References

1. Shaikh I, Küng SA, Aziz H, Sabir S, Shabbir G, Ahmed M, Dabash R. Telehealth for addressing sexual and reproductive health and rights needs during the COVID-19 pandemic and beyond: A hybrid t-community accompaniment model for abortion and contraception services in Pakistan. Front Glob Womens Health. 2021;2:705262.

2. Peng K, Tu K, Li Z, Hallinan CM, Laughlin A, Manski-Nankervis J-A, et al. Global impacts of COVID-19 pandemic on sexual and reproductive health services: An international comparative study on primary care from the INTRePID Consortium. BJOG. 2024;131(4):508–17.

3. Ng’andu M, Mesic A, Pry J, Mwamba C, Roff F, Chipungu J, et al. Sexual and reproductive health services during outbreaks, epidemics, and pandemics in sub-Saharan Africa: A literature scoping review. Systematic Reviews. 2022;11(1):161.

4. Nagendra G, Carnevale C, Neu N, Cohall A, Zucker J. The potential impact and availability of sexual health services during the COVID-19 pandemic. Sex Transm Dis. 2020;47(7):434–6.

5. Liu M, Zhou J, Lan Y, Zhang H, Wu M, Zhang X, et al. A neglected narrative in the COVID-19 pandemic: Epidemiological and clinical impacts of the COVID-19 outbreak on syphilis. Clin Cosmet Investig Dermatol. 2023;16:2485–96.

6. World Health Organization. HIV Switzerland, Geneva2022 [Available from: https://www.who.int/news-room/fact-sheets/detail/hiv-aids.

7. VanBenschoten H, Kuganantham H, Larsson EC, Endler M, Thorson A, Gemzell-Danielsson K, et al. Impact of the COVID-19 pandemic on access to and utilisation of services for sexual and reproductive health: a scoping review. BMJ global health. 2022;7(10):e009594.

8. Thoma ME, Declercq ER. All-cause maternal mortality in the US before vs during the COVID-19 pandemic. JAMA Network Open. 2022;5(6):e2219133-e.

9. Johnson KA, Burghardt NO, Tang EC, Long P, Plotzker R, Gilson D, et al. Measuring the impact of the COVID-19 pandemic on sexually transmitted diseases public health surveillance and program operations in the State of California. Sex Transm Dis. 2021;48(8).

10. Hollander JE, Carr BG. Virtually perfect? Telemedicine for Covid-19. N Engl J of Med. 2020;382(18):1679–81.

11. Lindberg LD, VandeVusse A, Mueller J, Kirstein M. Early impacts of the COVID-19 pandemic: findings from the 2020 Guttmacher survey of reproductive health experiences. New York: Guttmacher Institute, 2020.

12. Alexander GC, Tajanlangit M, Heyward J, Mansour O, Qato DM, Stafford RS. Use and content of primary care office-based vs telemedicine care visits during the COVID-19 pandemic in the US. JAMA Network Open. 2020;3(10):e2021476-e.

13. Centers for Medicare & Medicaid Services. Telemedicine 2020 [cited 2023 January 09]. Available from: https://www.medicaid.gov/medicaid/benefits/telemedicine/index.html.

14. Fatehi F, Wootton R. Telemedicine, telehealth or e-health? A bibliometric analysis of the trends in the use of these terms. J Telemed and Telecare. 2012;18(8):460–4.

15. Eysenbach G. What is e-health? J Med Internet Res. 2001;3(2):E20.

16. Lurie N, Carr BG. The role of telehealth in the medical response to disasters. JAMA Intern Med. 2018;178(6):745–6.

17. U.S. Department of Health and Human Services. Telehealth for chronic conditions 2022 [Available from: https://telehealth.hhs.gov/providers/telehealth-for-chronic-conditions/managing-chronic-conditions-through-telehealth/.

18. Hatef E, Wilson RF, Hannum SM, Zhang A, Kharrazi H, Weiner JP, et al. Use of telehealth during the COVID-19 Era. Systematic review. Rockville, MD: Agency for Healthcare Research and Quality, Johns Hopkins University Evidence-based Practice Center under Contract No.75Q80120D00003; 2022. Contract No.: 23-EHC005.

19. Yelverton V, Qiao S, Weissman S, Olatosi B, Li X. Telehealth for HIV care services in South Carolina: Utilization, barriers, and promotion strategies during the COVID-19 pandemic. AIDS Behav. 2021;25(12):3909–21.

20. LeVasseur B. Benefits of store and forward, or asynchronous telehealth solutions 2020 [Available from: https://hitconsultant.net/2020/02/24/benefits-of-store-and-forward-or-asynchronous-telehealth-solutions/.

21. Kampmeijer R, Pavlova M, Tambor M, Golinowska S, Groot W. The use of e-health and m-health tools in health promotion and primary prevention among older adults: a systematic literature review. BMC Health Serv Res. 2016;16 Suppl 5(Suppl 5):290.

22. Valenti WM. Sexual health in Rochester/Monroe County: Sexually transmitted infections are a shared problem with shared solutions. Trillium Health; 2021.

23. Elliott R, Timulak L. Why a generic approach to descriptive-interpretive qualitative research? Essentials of descriptive-interpretive qualitative research: A generic approach. Essentials of qualitative methods. Washington, DC, US: APA; 2021. p. 3–14.

24. Thompson Burdine J, Thorne S, Sandhu G. Interpretive description: A flexible qualitative methodology for medical education research. Med Educ. 2021;55(3):336–43.

25. Palinkas LA, Horwitz SM, Green CA, Wisdom JP, Duan N, Hoagwood K. Purposeful sampling for qualitative data collection and analysis in mixed method implementation research. Adm Policy Ment Health. 2015;42(5):533–44.

26. Hsieh HF, Shannon SE. Three approaches to qualitative content analysis. Qual Health Res. 2005;15(9):1277–88.

27. Polit DF, Beck CT. Nursing research: Generating and assessing evidence for nursing practice. 10th ed. Philadelphia, PA: Wolters Kluwer; 2017. 784 p.

28. Birks M, Chapman Y, Francis K. Memoing in qualitative research: Probing data and processes. Journal of Research in Nursing. 2008;13(1):68–75.

29. Miles MB, Huberman AM. Qualitative data analysis. 2nd ed. Thousand Oaks, CA: SAGE Publications; 1994. 338 p.

30. Coffey A, Atkinson P. Making sense ofqualitative data: Complementary research strategies. Thousand Oaks,CA: Sage.; 1996.

31. Patton MQ. Qualitative research and evaluation methods. Thousand Oaks, CA: Sage; 2002.

32. Morse JM, Field PA. Qualitative research methods for health professionals. 2nd ed. Thousand Oaks, CA: Sage; 1995.

33. Creswell JW, Poth CN. Qualitative inquiry research design: Choosing among five approaches. 4th ed. Los Angeles, CA: Diseases. 2020;47(7).

35. Andaya E, Bhatia R. Trading in harms: COVID-19 and sexual and reproductive health disparities during the first surge in New York state. Soc Sci & Med. 2023;339:116389.

36. Lunt A, Llewellyn C, Bayley J, Nadarzynski T. Sexual healthcare professionals’ views on the rapid provision of remote services at the beginning of COVID-19 pandemic: A mixed-methods study. Int J STD AIDS. 2021;32(12):1138–48.

37. Darlington CK, Hutson SP. Understanding HIV-related stigma among women in the Southern United States: A Literature Review. AIDS Behav. 2017;21(1):12–26.

38. Golub SA. PrEP stigma: Implicit and explicit drivers of disparity. Current HIV/AIDS Reports. 2018;15(2):190–7.

39. Simões D, Stengaard AR, Combs L, Raben D, partners TEC-iaco. Impact of the COVID-19 pandemic on testing services for HIV, viral hepatitis and sexually transmitted infections in the WHO European Region, March to August 2020. Eurosurveillance. 2020;25(47):2001943.

40. Zhang T, Mosier J, Subbian V. Identifying barriers to and opportunities for telehealth implementation amidst the COVID-19 pandemic by using a human factors approach: A leap into the future of health care delivery? JMIR Hum Factors. 2021;8(2):e24860.

41. Gustin TS, Kott K, Rutledge C. Telehealth etiquette training: A guideline for preparing interprofessional teams for successful encounters. Nurs Educ. 2020;45(2):88–92.

42. Rutledge CM, Kott K, Schweickert PA, Poston R, Fowler C, Haney TS. Telehealth and eHealth in nurse practitioner training: current perspectives. Adv Med Educ and Pract. 2017;8:399–409.

43. Ljungholm L, Edin-Liljegren A, Ekstedt M, Klinga C. What is needed for continuity of care and how can we achieve it? – Perceptions among multiprofessionals on the chronic care trajectory. BMC Health Serv Res. 2022;22(1):686.

44. Ohta R, Ikeda H, Sawa J. The importance of patient-centered care during the COVID-19 pandemic: lessons from a rural ward in Japan. J Rural Med. 2021;16(2):128–9.

45. Cuomo AM, Zucker HA, Dreslin S. Dear colleagues letter. Albany, NY: New York Department of Health AIDS Institute; 2020. p. 1–3.

46. Gunaratne SH, Hansen MB. Guidance: Adopting a patient-centered approach to sexual health. In: Institute NYSDoHA, editor. Clinical Guidelines Program. Baltimore, MD: Johns Hopkins University; 2022. p. 1–4.

47. Maier M, Samari G, Ostrowski J, Bencomo C, McGovern T. ’Scrambling to figure out what to do’: A mixed method analysis of COVID-19’s impact on sexual and reproductive health and rights in the United States. BMJ Sex Reprod Health. 2021;47(4):e16.

48. Thomas L, Capistrant G. State telemedicine paps analysis coverage and reimbursement Delray Beach, FL: American Telemedicine; 2016 [cited 2024 April 1]. Available from: https://mtelehealth.com/state-telemedicine-gaps-analysis-coverage-reimbursement/.

49. Shaver J. The state of telehealth before and after the COVID-19 pandemic. Prim Care. 2022;49(4):517–30.

50. Razavi S, Farrokhnia N, Davoody N. Nurses’ experience of using video consultation in a digital care setting and its impact on their workflow and communication. PLoS One. 2022;17(5):e0264876.

51. Ashcroft R, Sur, D., Greenblatt, A., & Donahue, P. The impact of the COVID-19 pandemic on social workers at the frontline: A survey of Canadian social workers. Br J of Soc Work, bcab158. 2021;bcab158.

52. Bern-Klug M, Beaulieu E. COVID-19 highlights the need for trained social workers in nursing homes. J Am Med Dir Assoc. 2020;21(7):970–2.

53. Booton RD, Fu G, MacGregor L, Li J, Ong JJ, Tucker JD, et al. Estimating the impact of disruptions due to COVID-19 on HIV transmission and control among men who have sex with men in China. medRxiv. 2020.

